# The Mental Health of Farm Wives

**DOI:** 10.64898/2026.07.20.26358460

**Authors:** Sharon May, R. M. Crossley

## Abstract

**Objectives:** Research on mental health in agriculture has increased in recent years; however, it remains largely focused on farmers themselves and is predominantly male-oriented. The mental health of farm wives and partners, many of whom play integral roles in farm operations, business management, and family life, remains difficult to characterise. This study therefore aims to explore the prevalence and causes of mental health challenges among farm wives and partners, and to investigate their use of, and barriers to, mental health support services.

**Methods:** Quantitative data was collected using over 450 structured questionnaire responses that assessed mental health prevalence, contributing stressors and support service utilisation.

**Results:** Findings indicate that there is a high prevalence of mental ill health amongst farm wives, seemingly due to industry stressors and support role overwhelm. Interpersonal relationships played a significant role in the types of mental distress experienced and highlighted the toll that farm life can take on farm wives’ social and emotional connections. Despite a range of formal and informal support services being available, and effective when used, significant barriers to accessing these services were identified, including both practical difficulties and self-stigmatisation due to cultural beliefs.

**Conclusions:** Farm wives and partners experience substantial mental health burdens linked to their diverse and often underrecognized roles within agricultural systems. In future, targeted interventions are needed to reduce stigma, improve service accessibility, and recognize women’s contributions within farm enterprises. Further research and dedicated investment are also essential to better understand and help improve the mental health of this overlooked population within agricultural industries.

## Introduction

Agriculture has historically been associated with demanding working conditions including long hours, physically intensive labour, and high levels of occupational stress [1]. Research consistently demonstrates that farmers experience poorer mental health outcomes than the general population, with elevated rates of suicide reported internationally [2]. In the UK, agricultural employees have suicide rates approximately 1.7–1.9 times higher than the national average, making farming one of the occupations with the greatest suicide risk [3].

The Royal Agricultural Benevolent Institution (RABI) facilitated the largest UK survey of health and wellbeing in agriculture involving over 15,000 participants and identified higher levels of mental ill-health compared to the wider population [4,5] . This “farmer wellbeing gap” is an area of considerable concern and led the UK Government Environment, Food and Rural Affairs Committee into Rural Mental Health to state that urgent and meaningful Government action on rural mental health is essential [6].

Whilst there are some nuances across nationalities and industry sectors, there are several common factors contributing to poor mental health in farming. Economic pressure driven by rising production costs, market instability, policy changes and uncertainty [4,7] is one of the most significant factors. Furthermore, agriculture depends heavily on weather conditions, and increasingly frequent extreme weather events threaten crop production, livestock welfare and financial sustainability [8,9]. Beyond this, farmers experience pressures associated with public narratives that attribute responsibility for climate change to agriculture [4]. As a result, compliance to ever-evolving regulatory requirements are also reported as a further contribution to psychological strain [4,10].

Isolation is another key contributor to poor emotional wellbeing, with almost a third of farmers hardly ever, or never, leaving the farm [4]. Rural location, declining local farming populations, and social and cultural disconnect leave almost half of farmers experiencing some level of loneliness [11,12].

Cultural expectations embedded within farming identity around independence and “rugged individualism” can discourage help-seeking behaviour, impact self-esteem and increase vulnerability when times are tough [10.13]. Long working hours and self-sacrifice are often culturally celebrated thereby normalising exhaustion and limiting opportunities for rest and support [12,14]. Physical injury, chronic pain and reduced mobility are also common in farmers—also closely linked to poor mental wellbeing and exacerbated by the culture expectation to keep pushing on [5,15].

In line with this stigma, agriculture is viewed as a traditionally masculine industry; however, women now account for approximately 27% of the paid UK agricultural workforce, and approximately 55% when considering unpaid work [16]. Whilst women have always played a significant role, their contributions to farming frequently remain undervalued and poorly captured in agricultural statistics [17]. The relative lack of research specifically into the mental health and wellbeing of women in farming means that there is very little understanding of any specific areas of need, which is imperative for designing and implementing effective interventions [18].

Rudolphi et al [19] found a similar proportion of high stress across both genders, particularly concerning personal finance and time. However, a higher proportion of women met the criteria for moderately severe and severe symptoms of anxiety and depression and experienced more significant isolation and loneliness, whereas a greater proportion of men fell into the mild category for anxiety and depression. Thus, whilst many of the beforementioned issues will affect the mental health and wellbeing of both genders, the differences in rural gender roles and expectations mean these issues can disproportionately impact women [18]. Conway and Mullane [20] previously identified five gender specific factors affecting the wellbeing of women in agriculture: gender-based discrimination, gender-biased mental health services, gender stereotypes, gender-related stigma regarding mental health and gender-specific help-seeking behaviours. For example, women who reported low support from a spouse or significant other were amongst the most stressed and were substantially more likely to report depression than men [19,21]

Further to this, the increasing economic pressures faced by farms have often required farm wives to diversify or seek additional income from off-farm employment without renegotiation of farm-related responsibilities [17,22]. While this can provide financial security, independence, and personal identity, it may also increase stress by adding further demands [23]. Rural gender norms continue to position motherhood and childcare as primarily women’s responsibility [16] in addition to community obligations, farm labour, and external employment [15,24]. These expectations create work overload and role conflicts, which are associated with poorer wellbeing and increased physical and psychological distress [23,25, 26], as well as negatively impacting farm productivity, profitability and safety, thereby further contributing to poor mental health and stress [21].

Unfortunately, ongoing struggles can regularly be compounded by feelings of guilt around not doing enough and finding difficulty in accessing support [21]. When it comes to help seeking, many women share the masculine stoicism and delay seeking support until distress becomes severe [25].

In order to understand how to help women farmers or farm wives better, we need to understand more about their experiences with mental ill health and accessing support for it. One challenge when understanding the picture of mental health of farm women is the variety of subsets of women working in agriculture: Women Farmers, Farm Wives, Farm Mothers, Farm Daughters, Female farm workers (including female migrant workers) and Entrepreneurial Women on Farm [22]. Studies that only look at farmers or farm workers risk excluding the experiences of other women on farm who don’t identify as such. Farm spouses are embedded in the household economy and exposed to many of the same physical and psychological stressors as farmers themselves regardless of whether they actively farm [15], and thus the wellbeing of farm wives is strongly influenced by the same pressures affecting their husbands [23]. There is also a similarity and symmetry to mental health within farming households where distress experienced by one partner frequently affects the other [14,23]. These ideas motivate the research presented in this study, which focuses on farm wives and partners of farmers as a specific subset of the women in the farming population. By understanding their needs and challenges in more detail, we hope to contribute to the growing research and provision for women in agriculture as a whole and ultimately help facilitate the development of effective support for this, regularly undervalued, subset of the agricultural community.

## Methods

The cross-sectional study used an anonymous online questionnaire to examine the prevalence of self-reported mental health difficulties among women who identified as wives or long-term partners of individuals working in agriculture. The study also sought to identify perceived stressors affecting mental health, patterns of support utilisation, and barriers to help-seeking.

Participants were recruited using convenience and snowball sampling methods. An advertisement containing a link to the online questionnaire was disseminated through social media and farming-related networks, with reminder posts circulated throughout the data collection period. The questionnaire included both closed and open-ended questions intended to explore participants’ experiences of mental health difficulties, their perceptions of the impact of farming life on mental health, the types of support services and resources accessed, and the perceived usefulness and barriers to accessing these support methods. A total of 466 participants completed the questionnaire and data were analysed descriptively using frequencies and percentages to summarise the results.

Ethical approval for the study was obtained from Bath Spa University in advance. Participation was voluntary, and informed consent was obtained electronically prior to data collection. Information about relevant and appropriate mental health support services was regularly provided throughout the study to support participant wellbeing.

## Results

Mental health difficulties were widely reported among the questionnaire’s respondents. Over 90% of individuals indicated experiencing mental health difficulties at some point in their lives, with 55% experiencing mental health issues at the point of study or within the last 12 months.

Anxiety was the most frequently self-reported condition, affecting 68.7% of respondents, followed by depression (53.9%) and grief or loss (30.5%) (Figure 1).

**Figure 1:**
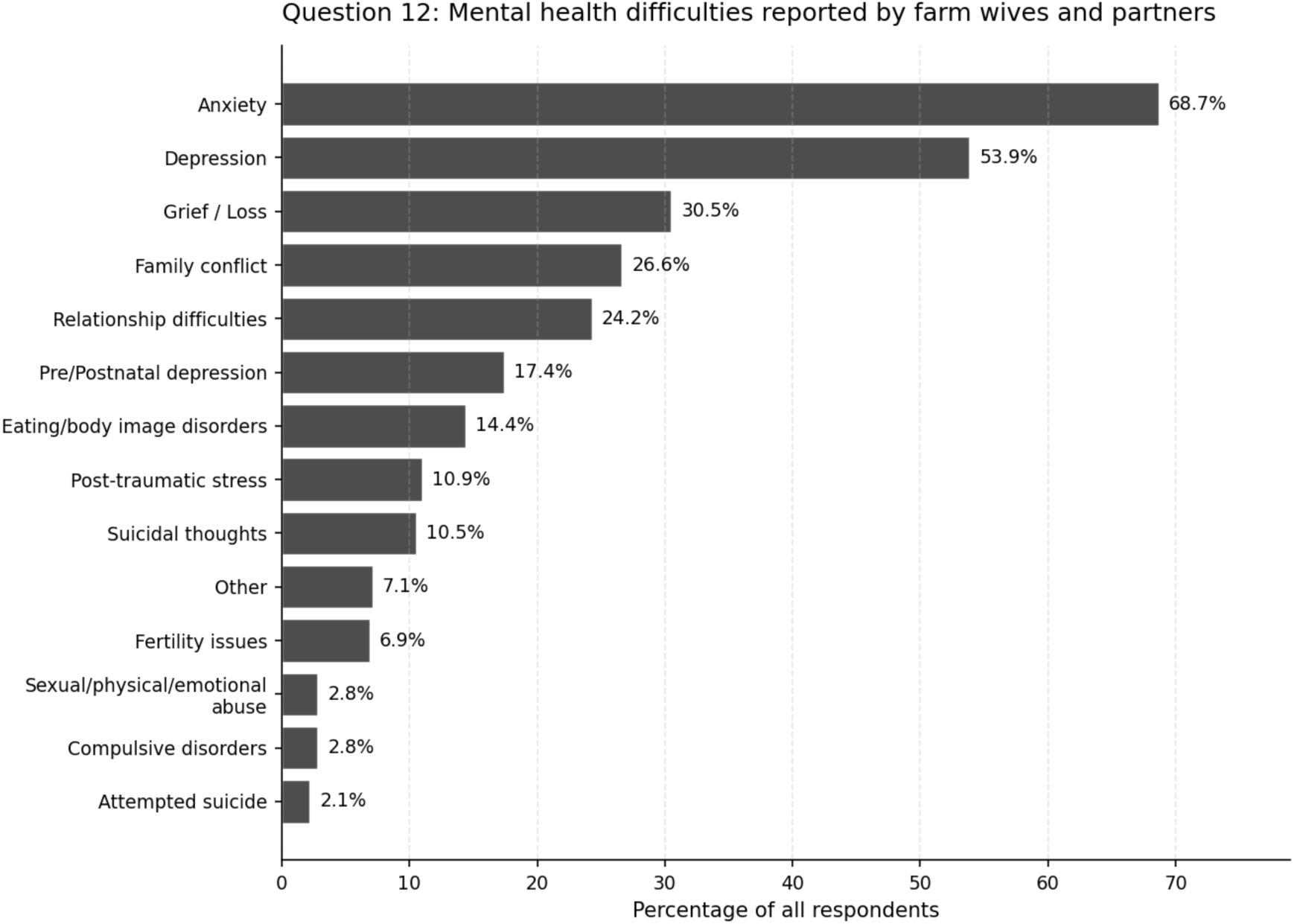
Summary of responses to questionnaire question 12, which asked “Would you categorise your struggles under any of the following headings? (tick all that apply)”.

Difficulties associated with family conflict (26.6%) and relationship difficulties (24.2%) were also commonly reported, highlighting the impact of interpersonal relationships on mental wellbeing. Smaller but notable proportions reported pre- or postnatal depression (17.4%), eating or body image disorders (14.4%), post-traumatic stress (10.9%), and suicidal thoughts (10.5%).

### The influences on the mental health of farm wives

Our findings demonstrate a substantial burden of psychological distress within the study population, extending beyond commonly recognised conditions such as anxiety and depression. Overall, 65% of respondents believed that working within the agricultural sector has negatively impacted their mental health. However, respondents also identified several aspects of farming life that supported their wellbeing, illustrating the complex and often contradictory nature of life in agriculture.

The most positively perceived aspects of farming were being outdoors and having access to the countryside, with almost all respondents rating these as mostly or totally positive influences on their mental health (Figure 2). Similarly, being around livestock and animals, raising children in a farming environment, and being part of the community were generally viewed as beneficial to wellbeing. These results are similar to previous findings that suggest that farming potentially has some protective characteristics for mental wellbeing which should be held in mind when considering the challenges [27].

**Figure 2:**
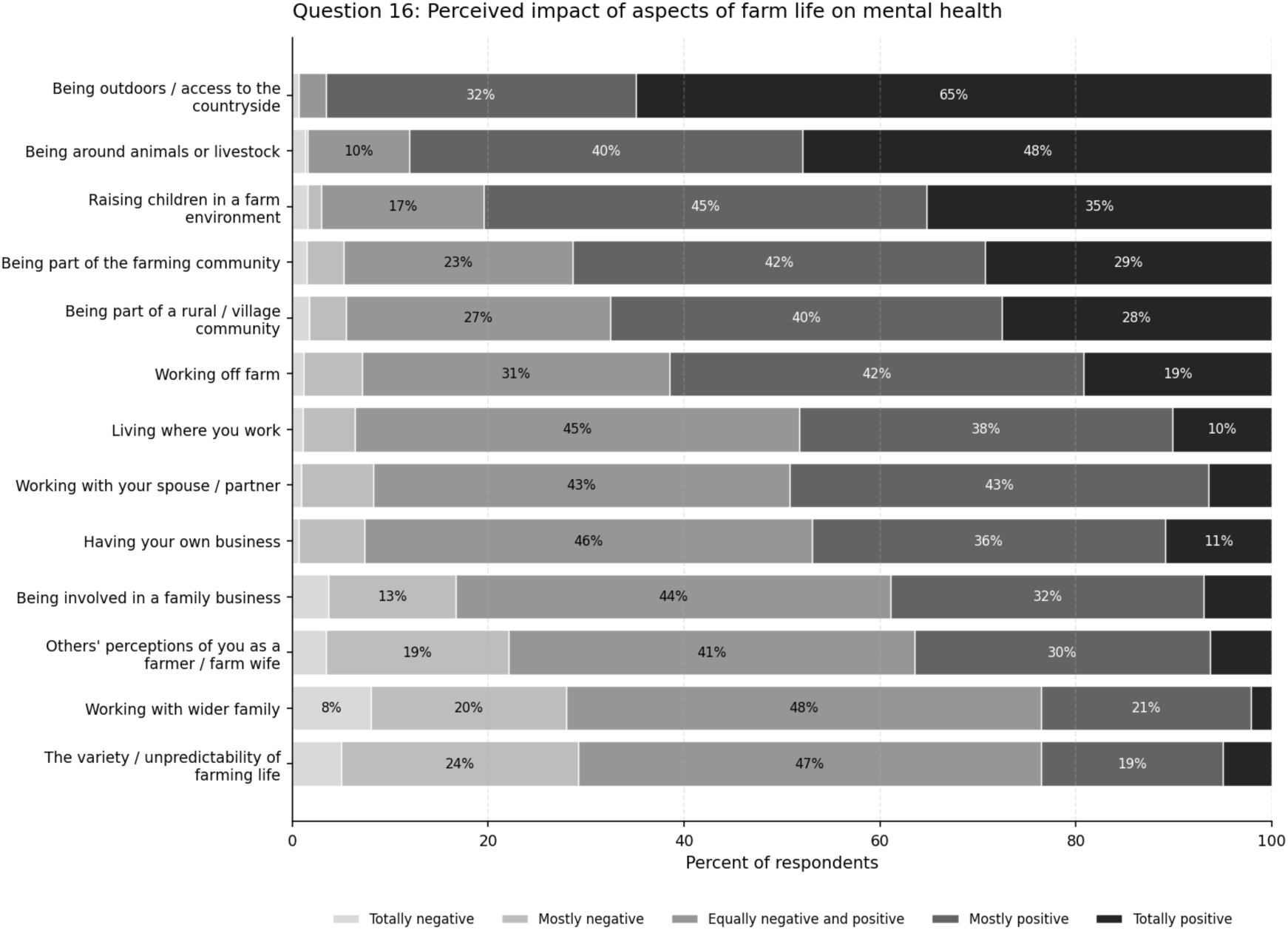
Summary of responses to questionnaire question 16, which asked “How would you categorise the impact of the following aspects of farm life on your mental health - please rank the aspects that apply to you and your situation.”.

Despite these positive aspects, respondents also identified substantial sources of stress associated with farming life (Figure 3). Financial pressure emerged as the most significant concern, with nearly two-thirds of respondents reporting that financial pressures frequently or constantly affected their mental health. Concerns about the future of the farm business and the wider farming industry were also commonly reported.

**Figure 3:**
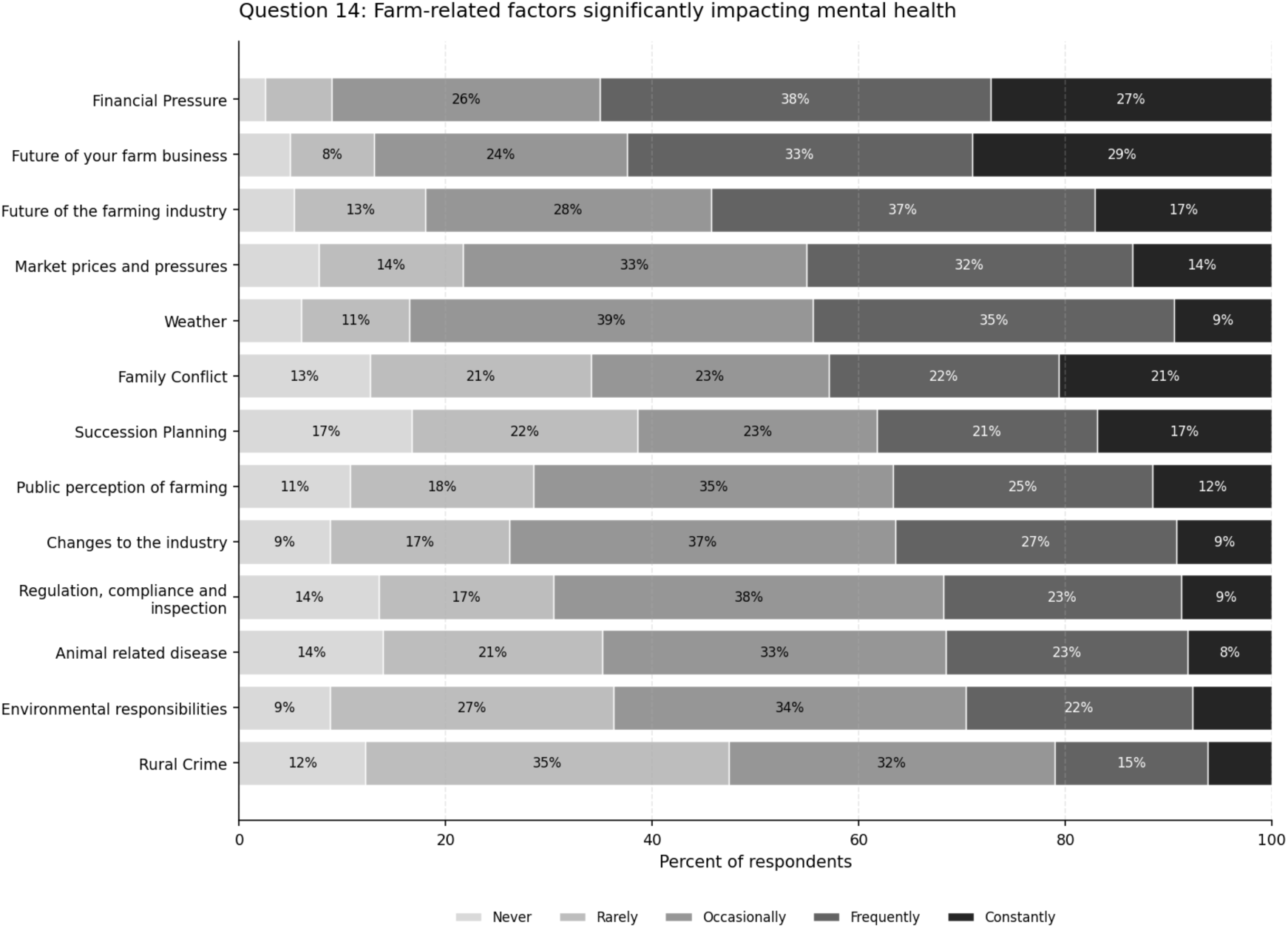
Summary of responses to questionnaire question 14, which asked “How often do you feel the following farm-related factors significantly impact your mental health?”.

Alongside these financial concerns, respondents frequently identified market volatility, weather-related uncertainty, family conflict, and succession planning as significant stressors. Regulatory requirements, public perceptions of farming, environmental responsibilities, and concerns relating to animal disease were generally reported as having a more moderate, but still important, impact on mental wellbeing. The wide-ranging list of stressors illustrates how agricultural industry demands extend far beyond the workplace and can impact those adjacent to, and not just, the principal farmer. One cause of this may be the close integration of home, work and family lives often found in these contexts. Whilst the stresses are similar to those of the RABI [4] survey there is a difference in the ranking order: we find financial issues being placed much higher than (for example) regulation and compliance compared to those findings. This is potentially due to the impact of the cost-of-living crisis that occurred in the UK between the two studies, whilst also the ongoing economic strain as a result of conflict in wider Europe.

Together, these findings suggest that respondents often viewed farming life as simultaneously rewarding and challenging. Many characteristics that attracted respondents to agricultural life remained sources of wellbeing, yet these coexisted alongside considerable economic uncertainty and occupational pressure that also negatively impacts an individual’s mental wellbeing.

### The role of farm wives is primarily one of supporting others

Respondents reported that the demands of farming had a substantial impact on family relationships and domestic life (Figure 4). The most frequently reported issue was that farming took priority over family needs, with more than three-quarters of respondents indicating that this occurred frequently or constantly. A lack of time together as a couple and as a family were also commonly reported consequences of the demands of farming life.

**Figure 4:**
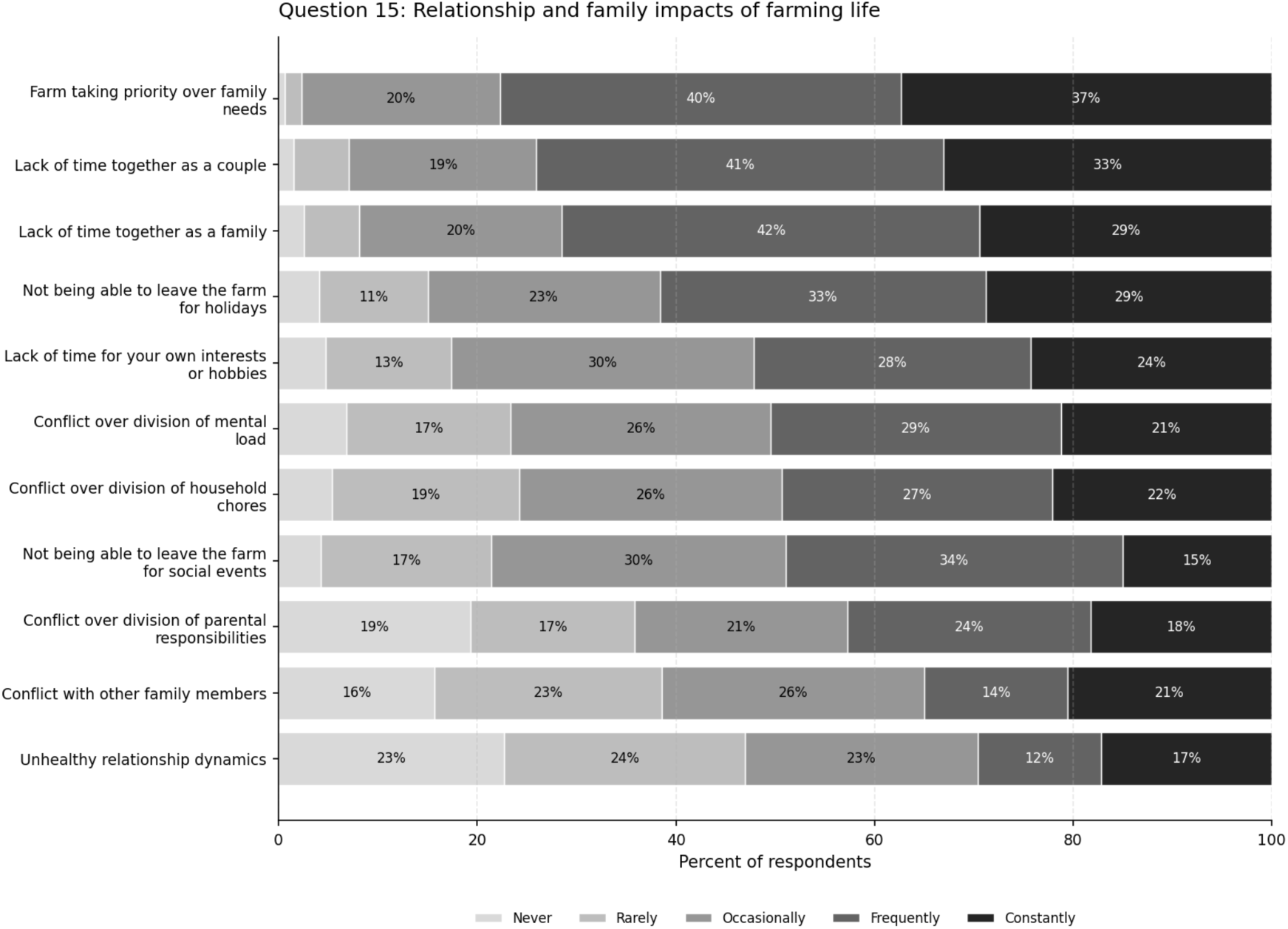
Summary of responses to question 15, which asked “Do you feel that being the wife or partner of a farmer results in the following? if so how often?”.

Although respondents generally valued being part of farming and rural communities (Figure 2), the demands of a farming lifestyle also limited opportunities for personal interests and social activities. More than half of respondents reported being unable to leave the farm for holidays or social activities on a regular basis, while many described having little time for hobbies or personal interests. These are all elements that signal potential decreased social and emotional connection and therefore increased isolation for these individuals: another potential contributor to poor mental wellbeing [11].

The complexity of respondents’ caring roles became particularly evident when considering the impact of supporting a partner experiencing mental health difficulties (Figure 5). Nearly three-quarters (73%) of respondents believed that their partner had experienced mental health difficulties. The most reported consequence was a negative effect on the respondents’ own mental health (41.2%), followed by relationship difficulties (37.3%) and increased feelings of loneliness or isolation (36.1%).

**Figure 5:**
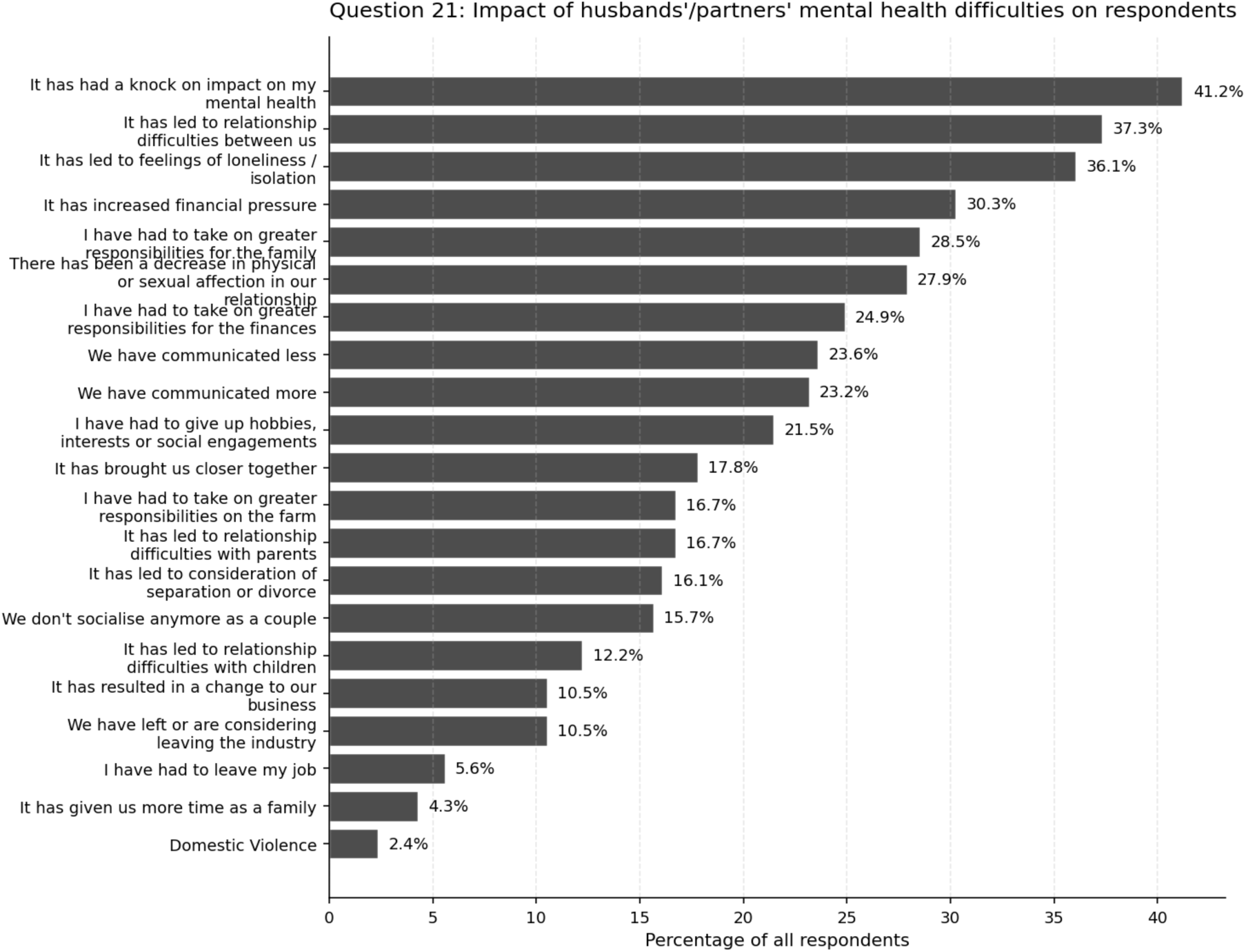
Summary of responses to questionnaire question 21, which asked “What impact has your husband / partner’s mental health struggle had on you? (tick any / all that apply)”.

Furthermore, many respondents also reported taking on additional responsibilities within the family, farm business, and household finances alongside their existing employment and caring responsibilities. Nearly one-third described increased financial pressure, while many reported assuming greater caring responsibilities when their partner was struggling. More than 15% had considered ending their relationship, while over 10% had left or considered leaving farming because of these challenges.

### Social support networks were perceived to help farm wives the most

Exploring the support utilisation of farm wives highlighted the importance of social connection as a source of support for maintaining their wellbeing (Figure 6). Informal support networks were consistently accessed more commonly than formal support services, indicating that relationships with spouses, friends, family members, and peers played a central role in supporting wellbeing. Among respondents who used them, informal support networks were perceived as highly beneficial. Close friends were frequently rated as providing useful or very useful support reinforcing the importance of those social connections for mental wellbeing [19].

**Figure 6:**
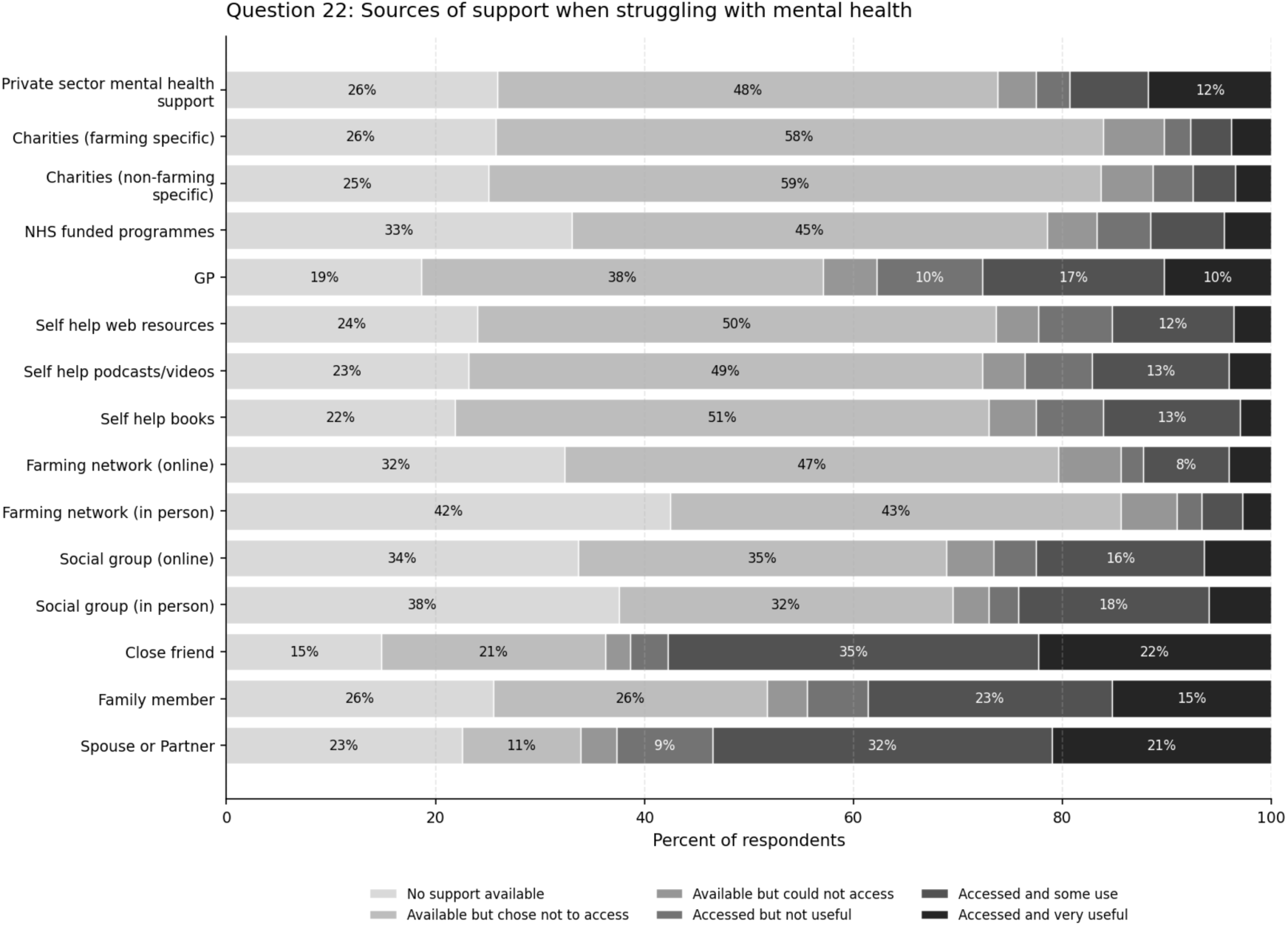
Summary of responses to questionnaire question 22, which asked “When struggling with your mental health, have you turned to any of the following for support?”.

Farming-specific support networks, both online and in-person, also appeared to play an important role. While fewer respondents accessed these services than informal family and friendship networks, those who did generally reported positive outcomes. This suggests that support delivered within farming contexts may provide benefits beyond traditional mental health services alone.

Formal services, including GPs, NHS-funded programmes, charities, counselling services, and private-sector mental health support, were accessed less frequently than informal sources of support. Nevertheless, respondents who engaged with these services generally reported positive experiences. Across all categories, many users reported that support had been either useful or very useful.

Overall, the findings indicate that support services were generally perceived positively once accessed, regardless of whether they were informal or formal sources of support. The primary challenge therefore appeared not to be the effectiveness of support itself, but rather whether individuals were able or willing to engage with available sources of help.

### Farm wives find it difficult to ask for support

The most commonly reported barrier to seeking support was a lack of time, affecting 62.0% of respondents. More than half also reported uncertainty about what type of support they needed, while 39.1% identified financial cost as a barrier to accessing help. The findings suggest that many barriers to accessing mental health support are interconnected. Time constraints may reflect the competing demands of farm work, childcare, off-farm employment, and household responsibilities. Similarly, financial concerns may relate not only to the direct cost of services but also to the indirect costs associated with travel, childcare, or taking time away from work.

One surprising result from this study included that despite widespread awareness of available support services, many respondents reported choosing not to access them (Figure 6). This pattern was observed across both formal and informal sources of support, suggesting that awareness alone did not necessarily translate into help-seeking.

Responses to the final open-ended survey question provided further insight into these barriers. A prominent theme was the identity of being the person who “holds everything together” or the individual that “everyone else turns to”, similar to that described in [13] as perceived masculinity. Many respondents described feeling responsible for supporting their partners, families, employees, and wider farming businesses while neglecting their own wellbeing, resulting in reluctance to admit they were struggling or to seek support themselves [21].

A second recurring theme was the lack of reciprocity in the support respondents received. Many described providing emotional, practical, and financial support to others while receiving little support in return, contributing to feelings of loneliness and emotional isolation. This finding relates closely to the results by Rudolphi [19] which found that women who reported low support from a significant other, or family member, were substantially more likely to report depression than men.

Finally, one significant consequence of this self-stigmatisation within this subset of females in agriculture was that respondents frequently described delaying help-seeking until they had reached a point of crisis or significant emotional distress [25]. Together, these findings demonstrate that barriers to accessing support extend beyond practical considerations such as time and cost, encompassing social expectations, caregiving responsibilities, and perceived cultural expectations to remain resilient.

## Discussion

This study provides a detailed examination of the mental health of farm wives and partners in the UK and highlights a substantial, yet often overlooked, burden of psychological distress within this population. While previous research has predominantly focused on the mental health of farmers themselves, our findings demonstrate that the psychological impact of agricultural life extends beyond the principal farmer to those who support farming businesses and families. Respondents described the cumulative impact of occupational, relational, and emotional pressures, illustrating that farm wives often experience multiple, interconnected stressors rather than a single source of psychological burden.

The high prevalence of self-reported mental health difficulties observed in this study is consistent with previous research demonstrating elevated levels of psychological distress within agricultural communities [4]. However, our findings extend this literature by showing that many respondents attributed their mental health difficulties directly to their involvement in farming, despite not necessarily identifying as the primary farmer. Furthermore, the mental health of respondents was influenced not only by the demands of agricultural work itself but also by the emotional consequences of supporting partners experiencing poor mental health. Together, these findings reinforce the importance of viewing mental health within farming households as a relational and family issue, rather than one affecting individual farmers in isolation.

A particularly important finding was the cumulative weight of the multiple roles undertaken by farm wives. Respondents frequently described balancing responsibilities within the farm business alongside off-farm employment, childcare, household management, financial administration, and emotional support for family members. Rather than identifying a single dominant stressor, participants described an ongoing accumulation of responsibilities that gradually eroded their own wellbeing. This pattern reflects the concept of role overload, whereby the sustained demands of multiple competing responsibilities contribute to chronic psychological strain. These findings also support previous research describing the often invisible and unpaid nature of women’s agricultural labour and the structural vulnerability that accompanies these roles within farming families [23]. Our qualitative findings suggest that the emotional labour undertaken by farm wives represents an equally important, yet frequently overlooked, component of agricultural work.

Although respondents identified many challenges associated with farming, they also recognised aspects of agricultural life that positively influenced their wellbeing. Access to the countryside, working with livestock, raising children within a farming environment, and belonging to rural communities were consistently viewed as important sources of satisfaction and resilience. These findings support emerging evidence that farming possesses characteristics that may protect mental wellbeing despite the considerable occupational challenges associated with the industry [27]. This apparent paradox highlights the complexity of farming life, where the same environment can simultaneously provide both psychological benefit and psychological strain.

The study also provides new insight into help-seeking among farm wives. Although practical barriers such as time pressures, financial constraints, childcare responsibilities, and geographical isolation were commonly identified, the qualitative findings suggest that these barriers represent only part of a more complex process. Many respondents described strong internalised expectations to remain resilient, prioritise the needs of others, and avoid appearing unable to cope. Several described themselves as the individual who “holds everything together” or the person that “everyone else turns to,” making it difficult to acknowledge their own needs or seek support. While previous studies have described the influence of masculine farming cultures on men’s help-seeking behaviours [13], our findings suggest that these cultural expectations may also be internalised by women living and working within agricultural environments.

Closely linked to this was the theme of reciprocity. Many respondents described providing practical, emotional, and financial support to their partners and families while perceiving little equivalent support in return. This imbalance frequently resulted in feelings of loneliness and emotional isolation despite living and working within close family units. These findings are consistent with previous research demonstrating that perceived social support is an important protective factor for mental wellbeing among women in agriculture [19]. Importantly, our findings indicate that the presence of social relationships alone may be insufficient; rather, the quality and reciprocity of those relationships appear central to psychological wellbeing.

Encouragingly, respondents generally viewed both informal and formal support services positively once they had been accessed. Informal networks, particularly family, friends, and farming peers, were most frequently utilised, while farming-specific support groups also received favourable evaluations. Formal mental health services, including GPs, counselling, charities, and NHS-funded services, were used less frequently but were similarly regarded as helpful by those who engaged with them. These findings suggest that the principal challenge may lie less in the perceived effectiveness of available support than in enabling individuals to access it before reaching crisis point. Interventions that focus solely on increasing service provision may therefore be insufficient without also addressing the practical, cultural, and psychological barriers that discourage help-seeking.

The findings have several implications for policy and practice. Mental health initiatives within agriculture have traditionally focused on farmers themselves; however, our findings suggest that farm wives and partners should be recognised as an equally important population for support. Their contribution extends beyond domestic responsibilities to include farm administration, business management, caregiving, and emotional support, all of which underpin the resilience of farming households. Interventions that adopt a family-centred approach, strengthen peer-support networks, improve flexibility of service delivery, and acknowledge the realities of agricultural life may therefore offer broader benefits than those targeting individual farmers alone. Equally important is the need to challenge cultural expectations surrounding caregiving, resilience, and self-reliance that may discourage women from seeking support for their own wellbeing.

Several limitations should be acknowledged. The use of convenience and snowball sampling may have introduced self-selection bias by attracting individuals with lived experience of mental health difficulties, while those without such experiences may have been under-represented. Mental health outcomes were self-reported and were not clinically validated. Although the survey included open-ended responses that enriched interpretation of the quantitative findings, more in-depth qualitative interviews would provide a richer understanding of the lived experiences of farm wives and partners. In addition, the framing of the survey may have unintentionally excluded more diverse family structures. Future research would benefit from longitudinal study designs, more diverse sampling strategies, and greater exploration of how mental health varies according to life stage, agricultural sector, employment status, motherhood, menopause, and experiences of suicidal ideation. Similar research should also be extended to other under-represented groups within agriculture.

In conclusion, this study provides further evidence that the mental health of farm wives and partners warrants greater recognition within agricultural, occupational health, and rural mental health policy. Although often not recognised as the primary farmer, these women occupy a pivotal role within farming households, balancing practical, emotional, and caregiving responsibilities while simultaneously navigating many of the same occupational pressures experienced by farmers themselves. Recognising and supporting this population is therefore likely to benefit not only individual wellbeing but also the resilience and sustainability of farming families more broadly. At the same time, respondents reminded us that farming is not defined solely by hardship. Many described deep satisfaction in the lifestyle, community, and natural environment associated with agriculture. Future research should therefore seek not only to reduce the factors that place farm wives at risk of poor mental health, but also to understand and strengthen the protective characteristics of farming life that respondents identified as central to their wellbeing.

## Disclosure

This research was undertaken whilst a student at Bath Spa University. No funding was provided, and no potential conflicts of interest were reported by the authors.

## Data Availability

All data produced are available online at https://github.com/beckycrossley/Data-MH-farm

## Acknowledgements

The authors would like to thank all the participants who gave their time to share their experience with us, and Roisin Ni Mhochain for her feedback and support.

## Notes

### Competing Interest Statement

The authors have declared no competing interest.

### Author Declarations

Ethics committee of Bath Spa University gave ethical approval for this work

